# BIOCHEMICAL AND HAEMATOLOGICAL CHANGES ASSOCIATED WITH TRANSPLACENTAL CONGENITAL MALARIA IN KOGI STATE, NORTHCENTRAL NIGERIA

**DOI:** 10.1101/2021.07.29.21261315

**Authors:** Shedrack Egbunu Akor, Dickson Achimugu Musa, SPO Akogu, Akpa Matthew, Emmanuel Titus Friday, Timothy Idachaba, Patience Onoja, Ameh Simon Abdul

## Abstract

**Background:** Transplacental congenital malaria is a vertical transplacental transmission of malaria parasites from the mother to the baby in utero or perinatally during labor. Cord blood that conveyed oxygen and nutrients from mother to fetus and return with carbon dioxide and other waste materials can transmit malaria pathogen. This study is aim to establish early diagnosis of transplacental congenital malaria using cord blood biochemical and haematological indices. Cord blood from 164 babies delivered at three hospitals in Kogi State between January and December, 2020 were microscopically investigated for malaria parasite. Biochemical and Haematological analyses were done using SYSMEX XP-300, Roche 9180 and VIS Spectrophotometer model 721. The data obtained were expressed as mean ±standard deviation using SPSS 23. The indicator level of statistical significance was set at p<0.05. The results showed significant (p<0.05) decreased in values of WBC, platelet, sodium, potassium, urea, creatinine, RBC, PCV, haemoglobin and MCHC in malaria infected cord blood in comparison to malaria negative control group. Significant (P<0.05) increased activities of liver enzymes (AST, ALT, ALP), total protein, bicarbonate and chloride in malaria infected cord blood when compared with malaria negative group. However, no statistically significant difference in lymphocyte, MCV, MCH, neutrophil and mixed of both malaria infected and malaria negative cord blood. This study suggests that cord blood biochemical and haematological indices can be used to diagnose and manage transplacental congenital malaria in fetus and neonates.

## Introduction

Malaria is one of the leading haemoparasite that causes diseases of human. The disease is caused by protozoa parasite of the genus plasmodium, which gets injected into the human bloodstream through the bite of an infected female Anopheles mosquito. Transplacental congenital malaria is a vertical transplacental transmission of malaria parasites from the mother to the baby in utero or perinatally during labor (1). Worldwide, it has been estimated that each year between 75,000 and 200,000 infant deaths are attributed to congenital malaria in pregnancy (2). Studies have shown before now that the rate of transplacental congenital malaria is rare in most of the endemic countries in sub-Saharan Africa and India compared to non-endemic regions like Europe and South East Asia (3). Plasmodium falciparum is the main specie of plasmodium that causes congenital malaria in endemic regions while Plasmodium vivax and Plasmodium malariae are the major causative agents in non-endemic areas. The prevalence of congenital malaria in both endemic and non-endemic areas are 0.3% and 46.7% respectively (2). In endemic countries, congenital malaria was formally considered to be a rare condition due to protective effect supplied by considerable immunity to malaria acquired through mothers’ passive maternally transferred antibodies on parasite and protective factors of the placenta (1, 4). Reports of recent studies have shown high prevalence of congenital malaria both in endemic and non-endemic regions which has been attributed to increased resistance to available antimalarial drugs and altered antigenic determinants of malaria parasites that causes parasitic virulence (5). Clinically, unlike signs and symptoms of malaria in older children, the symptoms of congenital malaria are non-specific and are often confused with neonatal sepsis syndromes. Though fever is the main symptoms of malaria but could be absent in congenital malaria as it symptoms overlap with the symptoms of neonatal sepsis (6). The clinical features of congenital malaria which most times overlap with neonatal sepsis include refusal to suck, anaemia, jaundice, diarrhoea, excessive crying, vomiting, lethargy, convulsions, irritability, tachypnoea, respiratory distress, hepatosplenomegaly (7). The postulated mechanisms of transplacental transmission of malaria parasites from mothers to the newborns comprises of different mechanisms which includes; direct chorionic villi penetration, maternal transfusion to fetal circulation due to premature separation of placenta at the time of delivery or rupture during pregnancy (8). In newborn babies, the progression of the malaria parasite from infection to clinical disease presentation is still cumbersome and not fully understood. Observations in some newborn babies with parasitemia at birth have shown spontaneous clearing of the parasite without antimalarial and the newborns do not develop clinical malaria (9). This remarkable protective capability of babies to resist malarial infection has been attributed to transmission of immunoglobulin G antibodies (IgG) against malaria to fetus from mothers that acquired considerable immunity which confers transient protection to the infants (10). Increased level of fetal haemoglobin (HbF) in the newborns is also a known factor that slows down the rate of malaria parasite development in neonates (11). With age, the protective ability of neonates to malaria parasites decreases due to diminished levels of infants’ HbF and maternal IgG.

The umbilical cord is the conduit pipe like structure that connects fetus at the abdomen with the placenta and serves as lifeline between the fetus and the mother (12). The umbilical vein and arteries convey oxygen and nutrients from mother to fetus and return with carbon dioxide and other waste materials from the fetus respectively (13, 14). Damages caused to umbilical cord by either microorganisms or chemical toxins are likely to reflect on the wellbeing of the fetus (12). Plasma, platelets, red blood cells and white blood cells are present in the Cord blood as contained in the adult whole blood which can be altered by malaria parasites (15). In addition, the umbilical cord contains biomolecules and electrolytes, such as proteins, lipids, carbohydrates, nucleic acids/nucleotides, enzymes, chloride, sodium, bicarbonate, potassium and trace of uric acid (16). To maximize the chances of early diagnosis and treatment of congenital malaria, use of umbilical cord blood is important. Therefore, this study aims to institute early diagnosis of congenital malaria using cord blood to evaluate the effects of malaria parasites on neonates’ biochemistry and haematology.

## MATERIALS AND METHOD

### Study design and subjects

This study is a cross-sectional research that involved newborn babies whose umbilical cord blood biochemical and haematological parameters were assessed for biomarkers of transplacental congenital malaria. A total of one hundred and sixty four [164] participated babies’ cord blood delivered at Kogi State University Teaching Hospital, Grimard Catholic Hospital and Amazing Grace Hospital were enrolled into the study. The study was approved by Grimard Catholic Hospital, Amazing Grace Hospital and the ethical clearance committee of Kogi State University Teaching Hospital. Cord blood samples and data were collected between February and December, 2020. All the participants’ parent/guardian in this study gave their informed consent. This was done via an informed consent form duly completed by all the subjects Parent/Guardian.

### Sample size determination

The calculation of minimum sample size was obtained based on the population size of 285 pregnant women registered for antenatal care and were due for delivery at these hospitals during the period of this study. 95% confidence level, 5% margin of error and population proportion of 50% were considered to have sample size of 164 participants needed to have a confidence level of 95% that the real values were within ±5% of the measured values.

### Sample Collection

4.5 mL of arterial cord blood was collected into a 5mL specimen EDTA collection tubes. The EDTA tube was inverted 8 times. 5ml of arterial cord blood was also collected and dispensed into plain sample container, allowed to clot and retract. The serum was separated from the clotted blood after centrifugation using Eppendorf Centrifuge 5804R at 27 °c and stored in an ultra-freezer at -80°c within 30minutes of separation.

### Thick and thin film for malaria microscopy

Thick and thin cord blood films were prepared immediately upon cord blood collection on the same slide. For thick film, 12 microlitre of cord blood was spread over a diameter of 15mm, while 2 microlitre of cord blood was used for thin films. The thin film was then fixed in absolute methanol for 2 seconds and air-dried. The cord blood films were stained after 24 hours with 3% Geimsa stain solution at pH 7.2. The air-dried stained slides were viewed under the microscope at X100 oil emersion and malarial parasites densities were counted per high power field (hpf) as follows: 1 parasite per hpf=Low density (+), 2-9 parasites per hpf=medium density (++), >20 parasites per hpf=high density (+++).

### Haematological assay

Automated SYSMEX XP-300 haematology analyzer was used to analysed cord blood haematological parameters within two hours of collection. This automated haematology analyzer is a three parts differential machine that gives differential percentage results of neutrophil, lymphocyte and mixed (monocyte, eosinophil, basophil) and the count values of white blood cell, red blood cell, haemoglobin, packed cell volume, mean cell volume, platelet, mean cell haemoglobin and mean cell haemoglobin concentration.

### Biochemical assay

Photometric determination of serum creatinine and urea were analysed using Randox diagnostic kits according to the method of Veniamin and Vakirtzi-Lemonias 1970 (17) and Tietz et al., 1994 (18) respectively. Serum total protein was carried out using biuret method, serum AST and ALT were colorimetrically determined according to Reitman and Frankel method 1957 (19), ALP was analyzed using method as described by Quimica Clinica Aplicada (QCA) S.A while serum electrolytes were analysed using Roche 9180 automated electrolytes analyser and Bicarbonate done colorimetrically using TECO reagent. All the analyses were done at Medical Laboratory Department, Kogi State University Teaching hospital, Anyigba, Kogi State.

### Statistical analysis

The data obtained were expressed as mean ± standard deviation using one way ANOVA of SPSS statistical software, version 23.0. The indicator level of statistical significance was set at P<0.05

## RESULTS

A total of one hundred and sixty four [164] participated babies’ cord blood delivered were enrolled into the study. Forty four (26.9%) babies’ cord blood had *P.falciparum* malaria while one hundred and twenty (73.2%) were negative and served as control group. Of the babies’ cord blood with malaria, 26 (15.9%) had + degree of parasitemia while 18 (11.0%) were ++ degree of parasitemia (Table 1). Table 2 shows the mean value of the haematological parameters in both malaria parasitemic and negative controls cord blood. There was significant (P<0.05) reduction in the WBC, RBC, haemoglobin, PCV, MCHC and platelet levels in cord blood with *P.falciparum* compared to the control group. However, no significant difference in MCV, MCH, neutrophil, lymphocyte and mixed (monocyte, eosinophil, and basophil) counts of the malaria positive cord blood when compared with malaria negative cord blood. The mean count of platelet was significantly (P<0.05) lower in cord blood with ++ degree of malaria parasitemia compared to + while MCH level in the cord blood with + malaria parasitemia was significantly higher than the cord blood with ++ degree of malaria parasitemia (P<0.05). Figure 1 shows the graphical presentation of mean count values of haematological parameters and babies’ cord malaria status.

**TABLE 1:**
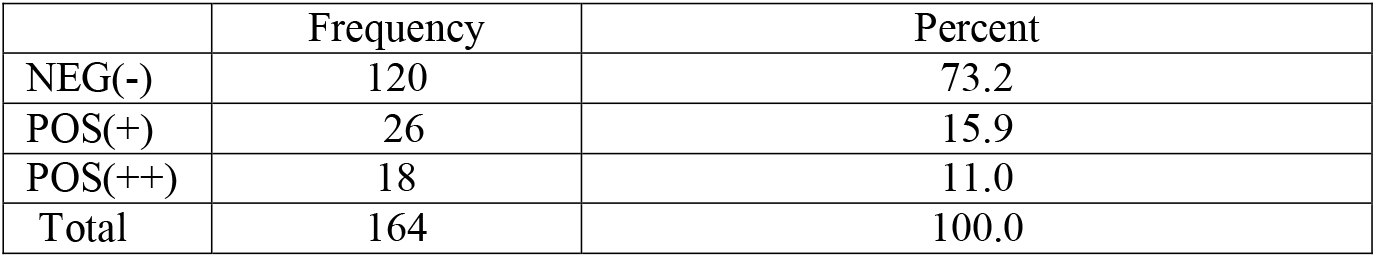
Frequency Distributions of babies Cord Blood Malaria Status.

**Table 2:** Mean count of haematological parameters associated with malaria status of babies cord blood.

| Parameters | Pos. (+)<br>(n=26) | Pos. (++)<br>(n=18) | Neg.(-)<br>(n=120) | ANOVA<br>(F) |
| --- | --- | --- | --- | --- |
| WBC ($\times 10^9/\text{L}$) | $8.0 \pm 3.7^*$ | $8.3 \pm 6.1^*$ | $11.7 \pm 5.7$ | 6.934 |
| RBC ($\times 10^{12}/\text{L}$) | $2.7 \pm 1.3^*$ | $2.9 \pm 1.0^*$ | $4.0 \pm 1.0$ | 19.067 |
| Haemoglobin (g/dl) | $8.5 \pm 3.1^*$ | $9.9 \pm 3.5^*$ | $13.1 \pm 2.7$ | 34.5 |
| PCV (%) | $28.4 \pm 10.0^*$ | $32.9 \pm 12.2^*$ | $42.7 \pm 9.3$ | 27.194 |
| MCV (fl) | $112.3 \pm 13.2$ | $113.2 \pm 8.6$ | $110.1 \pm 13.1$ | 0.667 |
| MCH (pg) | $31.8 \pm 3.4$ | $33.4 \pm 2.4$ | $34.0 \pm 6.6$ | 1.455 |
| MCHC (g/dl) | $28.4 \pm 1.5^{**}$ | $29.5 \pm 1.0^*$ | $30.4 \pm 2.5$ | 9.836 |
| Platelets ($\times 10^9/\text{L}$) | $106.7 \pm 85.0^*$ | $56.2 \pm 43.2^{**}$ | $147.8 \pm 79.1$ | 12.614 |
| Neutrophil (%) | 49.1±12.4 | 54.1± 12.1 | 49.8±17.2 | 0.708 |
| Lymphocyte (%) | 37.7±9.6 | 34.9±9.3 | 35.2±14.1 | 0.400 |
| Mixed (%) | 13.6±8.2 | 11.1±4.1 | 15.4±14.53 | 0.962 |

WBC: total white cell count; RBC: red blood cell; PCV: packed cell volume, MCV: mean cell volume; MCH: mean cell haemoglobin; MCHC: mean cell haemoglobin concentration, MIXED: (monocyte, eosinophil, and basophil).
Values expressed as mean ± S.D. \* and mean ± S.D. \*\* are significantly ( $P<0.05$ ) different from malaria negative group (Neg.-) while values expressed as mean ± S.D. \*\* only shows the level of significant ( $P<0.05$ ) different with respect to degree of parasitemia between malaria parasite positive (Pos. +) and positive (Pos. ++) of babies cord blood sample

**Figure 1:**
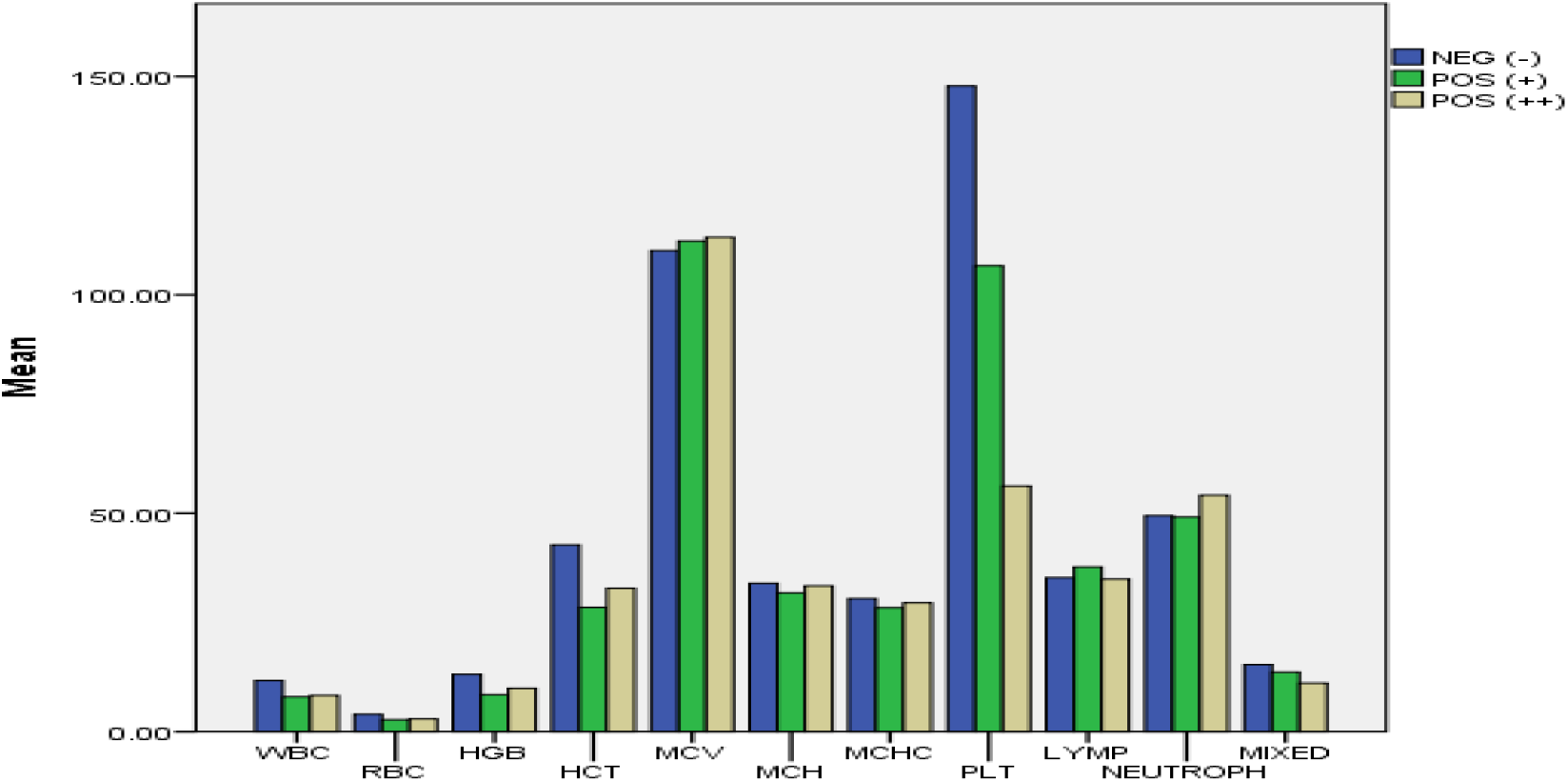
Mean count of WBC: Total White Cell Count; RBC: Red Blood Cell; PCV: Packed Cell Volume, MCV: Mean Cell Volume; MCH: Mean Cell Haemoglobin; MCHC: Mean Cell Haemoglobin Concentration, Mixed: (monocyte, eosinophil, basophil)

The effects of *p. falciparum* on cord blood biochemistry was also carried out and the mean count values results of biochemical parameters analysed are presented in table 3 and figure 2. The biochemical analyses results showed significant increase (P<0.05) in the ALP, AST, ALT, Total protein, Bicarbonate (HCO_3_^-^) and Chloride (Cl^-^) of the malaria infected cord blood compared to malaria negative cord blood, significant decrease (P<0.05) in the Urea, Sodium (Na^2+^), Potassium (K^+^) and Creatinine of the malaria infected cord blood compared to non-infected cord blood. The activities of ALP, AST, ALT, Total protein, Bicarbonate (HCO_3_^-^) and Chloride (Cl^-^) were significantly (P<0.05) higher in cord blood with ++ degree of malaria infection compared to + degree malaria infected cord blood while the mean count levels of Urea, Sodium (Na^2+^), Potassium (K^+^) and Creatinine of the + degree malaria infected cord blood had significant (P<0.05) higher levels when compared with ++ malaria infected cord blood.

**Table 3:**
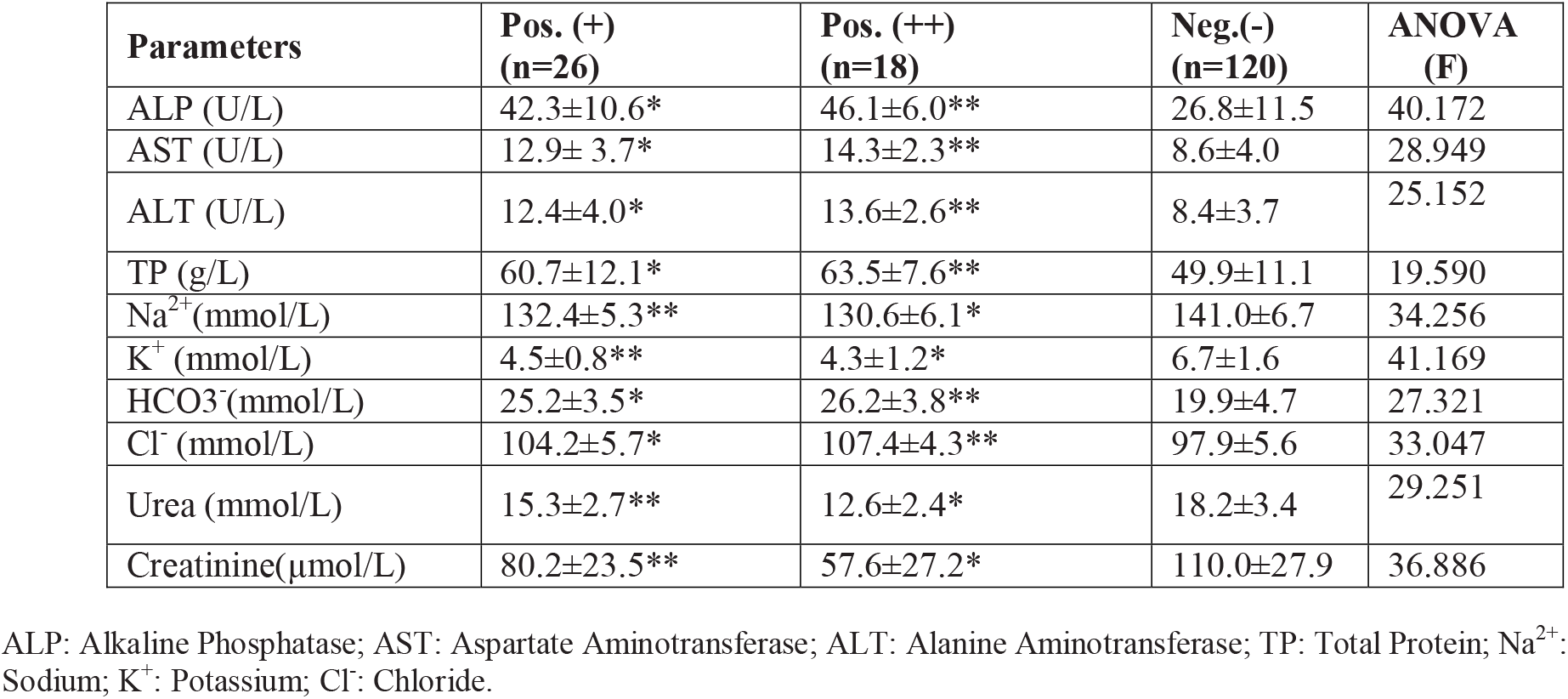
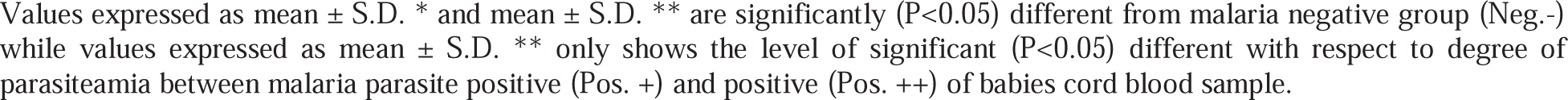
Mean count of biochemical parameters associated with malaria status of babies cord blood.

| Parameters | Pos. (+)<br>(n=26) | Pos. (++)<br>(n=18) | Neg.(-)<br>(n=120) | ANOVA<br>(F) |
| --- | --- | --- | --- | --- |
| ALP (U/L) | 42.3±10.6* | 46.1±6.0** | 26.8±11.5 | 40.172 |
| AST (U/L) | 12.9± 3.7* | 14.3±2.3** | 8.6±4.0 | 28.949 |
| ALT (U/L) | 12.4±4.0* | 13.6±2.6** | 8.4±3.7 | 25.152 |
| TP (g/L) | 60.7±12.1* | 63.5±7.6** | 49.9±11.1 | 19.590 |
| Na<sup>2+</sup>(mmol/L) | 132.4±5.3** | 130.6±6.1* | 141.0±6.7 | 34.256 |
| K<sup>+</sup> (mmol/L) | 4.5±0.8** | 4.3±1.2* | 6.7±1.6 | 41.169 |
| HCO<sub>3</sub><sup>-</sup>(mmol/L) | 25.2±3.5* | 26.2±3.8** | 19.9±4.7 | 27.321 |
| Cl<sup>-</sup> (mmol/L) | 104.2±5.7* | 107.4±4.3** | 97.9±5.6 | 33.047 |
| Urea (mmol/L) | 15.3±2.7** | 12.6±2.4* | 18.2±3.4 | 29.251 |
| Creatinine(μmol/L) | 80.2±23.5** | 57.6±27.2* | 110.0±27.9 | 36.886 |

ALP: Alkaline Phosphatase; AST: Aspartate Aminotransferase; ALT: Alanine Aminotransferase; TP: Total Protein; Na<sup>2+</sup>: Sodium; K<sup>+</sup>: Potassium; Cl<sup>-</sup>: Chloride.
Values expressed as mean ± S.D. \* and mean ± S.D. \*\* are significantly ( $P<0.05$ ) different from malaria negative group (Neg.-) while values expressed as mean ± S.D. \*\* only shows the level of significant ( $P<0.05$ ) different with respect to degree of parasitemia between malaria parasite positive (Pos. +) and positive (Pos. ++ of babies cord blood sample.

**Figure 2:**
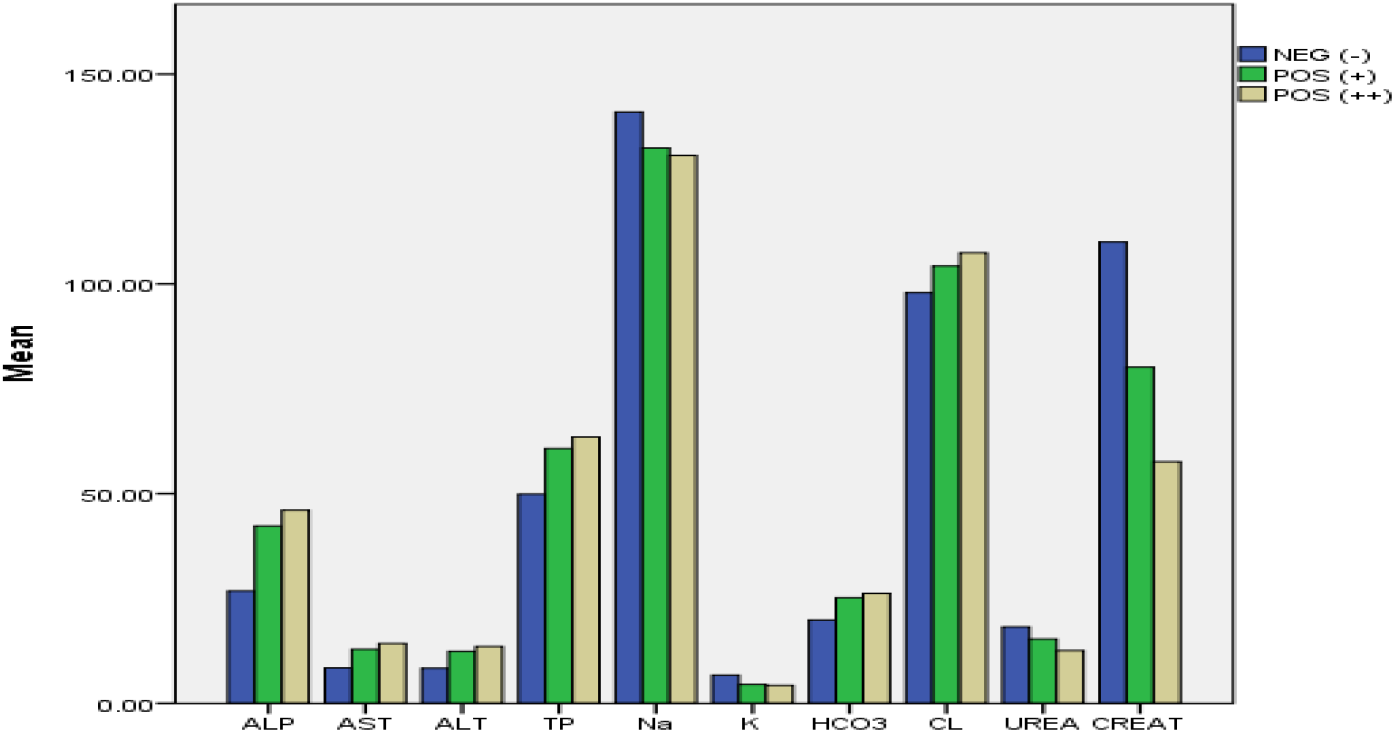
Mean count of ALP: Alkaline Phosphatase; AST: Aspartate Aminotransferase; ALT: Alanine Aminotransferase; TP: Total Protein; Na^2+^: Sodium; K^+^: Potassium; Cl^-^: Chloride; HCO3: Bicarbonate.

## Discussion

Biochemical and Haematological parameters are clinically used to evaluate individual health status. Early diagnosis and management of transplacental congenital malaria in fetus and neonates is a public health problem in sub-Saharan regions owing to multiple clinical signs and symptoms involved. The current study put in a well-designed clinical and laboratory procedures that uses umbilical cord blood biochemical and haematological indices to establish early diagnosis and management of transplacental congenital malaria in fetus and neonates.

Cord blood haematological indices obtained from this study may be useful in early diagnosis of transplacental congenital malaria as associated changes were observed when compared with negative control group. The present study showed significant (p<0.05) decrease in mean count values of white blood cells (leucopenia), platelet (thrombocytopenia), red blood cell (anaemia) and other red blood cell indices such as packed cell volume, haemoglobin and mean cell haemoglobin concentration in malaria infected cord blood in comparison to malaria negative control group (Table 2). The significantly (p<0.005) decreased mean count value of WBC observed among cord blood of babies infected with malaria compared to malaria negative control in this study is in consonance with previous reports of characterized leucopenia during malaria infection (20, 21, 22). But this finding is at variance with previous report from India where leucocytosis was reported among children infected with malaria (23) However, several researchers suggested that leucocytosis obtained from these reports may be associated with poor prognosis or concurrent malaria infection (24, 25, 26). The mechanism of leucopenia in malaria infection is still a topic of research but thought have suggested that it may be due to splenic sequestration, leucocytes localization away from circulatory blood and other marginal pools rather than actual stasis (27).

Anaemia and thrombocytopenia associated with malaria infection in neonates have been widely reported as sensitive and specific biomarkers for malaria diagnosis in babies (28). This present study observed significant (p<0.005) reduction in mean count value of RBC, PCV, MCHC and haemoglobin suggestive of malaria associated anaemia among malaria infected cord blood compared to malaria negative cord blood. The pathogenicity of anaemia associated with malaria in neonate still remains complex as multi-factors are involved. Multiplication of malaria parasite at different stages of development may results in rupture of cord RBCs and subsequent decrease in the level of PCV, MCHC and haemoglobin. Secondly, decreased erythropoiesis activity and shortened life span of infected RBCs are also part of this postulate (29, 30) However; the severity of anaemia associated with malaria infection in this finding did not correlate with the degree of parasitaemia. This report concurred to that of Karolina et al (31).

In addition to anaemia associated with malaria observed among cord blood infected with malaria parasite, this study also observed low platelet count (thrombocytopenia) among cord blood infected with *P. falciparum*. The postulated mechanisms of thrombocytopenia associated with malaria infection are still cumbersome. *P. falciparum* induced peripheral platelet destruction and platelet consumption by disseminated intravascular coagulation (DIC) are the major suggestions reported (32, 33, 34).

Biochemical data obtained from this study indicate significant increased (P<0.05) mean value of alkaline phosphatase (ALP) in babies cord blood infected with malaria when compared with malaria negative cord blood. The significant increased levels of ALP in malaria infected cord blood could be an indication of defensive mechanism role ALP plays in defending the umbilical cord against vertical transmission of malaria pathogen from mother to fetus (35). The role of ALP during malaria infection could be as a result of polymorphonucleated (PMN) cells and granulocyte colony-stimulating factor activations cascades during infections that causes increased in serum ALP which is in accordance with the previous reports (36). However, because of the bone formation process in neonates, high levels of serum ALP activity in newborn cord blood may not be a useful biomarker of transplacental congenital malaria infection in children (37). AST and ALT results obtained in this study showed the degree of significant (P<0.05) between malaria infected and malaria negative cord bloods, the activities of AST and ALT in malaria infected cord blood were significantly higher compared to the activities in non-malaria infected cord blood. This increased activities of liver enzymes observed among malaria infected cord blood in this study could demonstrate leakage of transaminases and alkaline phosphatase from parenchymal and hepatocytes membrane respectively into the circulatory system or exchanged cord blood between the mother and the child as reported previously (38, 39). According to this present study, the total protein concentration in malaria infected cord blood is statistically significance (P<0.05) when compared to malaria negative cord blood total protein. This may indicates the role of certain proteins in the cord blood as the body fights malaria infection or other inflammation (40). The degree of malaria parasiteamia significantly altered these biochemical parameter; serum ALP, AST, ALT and total protein show higher mean values in ++ degree of parasiteamia when compared with that of a +. This indicates positive correlations between the degree of cord blood malaria infection and the levels of these parameters in the cord blood.

Serum blood urea nitrogen (BUN) and creatinine are the routinely used biomarkers that provide useful information concerning the health status of the renal system. The mean values of cord blood urea and creatinine obtained in this study were significantly (P<0.05) higher among malaria negative when compared to malaria infected cord blood samples. Although the mechanism behind this is still a topic for further study. This finding is in accord with the previous report of significantly higher level of urea and creatinine in cord blood samples not infected with P. falciparum when compared to cord blood infected by P. falciparum (36, 41).

Despite significant (p<0.005) discrepancies observed with analyzed electrolytes (sodium, potassium, Bicarbonate and chloride) in this study between malaria infected and malaria negative cord blood. The mean ± SD of sodium, potassium, Bicarbonate and chloride of both malaria infected (+, ++) and malaria negative cord blood obtained are within the normal reference values for each. These findings again correspond with other studies done before now where normal values of electrolytes were recorded for both infected and non-infected neonates (42).

### Conclusion

This study suggest that presence of cord blood leucopenia, thrombocytopenia, anaemia, high activities of liver enzymes and total protein with decreased mean count values of sodium, potassium, urea, creatinine and significant increased values of bicarbonate and chloride of cord blood at birth increases the probability of transplacental congenital malaria that needs prompt clinical management and treatment to reduce neonatal morbidity and mortality. Therefore early diagnosis and treatment of congenital malaria using cord blood biochemical and Haematological indices is advocated

### Recommendation

This report looks at malaria parasite alone. It will be important to further investigate the effect of other neonatal pathogens on cord blood biochemical and haematological parameters in next studies.

## Data Availability

Availability of data and materials: The study data is available on personal request to the corresponding author.

## Acknowledgements

The authors wish to acknowledge the staff of Kogi State University Teaching Hospital, Grimard Catholic Hospital and Amazing Grace Hospital. Especially the Medical Doctors, Nurses and Medical Laboratory Scientists for their various contributions to this work.

## Declaration

Author(s) declared that this study was not funded and no competing interest

## Availability of data and materials

The study data is available on personal request to the corresponding author.

## Notes

### Competing Interest Statement

The authors have declared no competing interest.

### Author Declarations

The study was approved by Grimard Catholic Hospital. Amazing Grace Hospital and the ethical clearance committee of Kogi State University Teaching Hospital. All the participants parent/guardian in this study gave their informed consent. This was done via an informed consent form duly completed by all the subjects Parent/Guardian.

